# Effects of SARS-CoV-2 Alpha, Beta, and Delta variants, age, vaccination, and prior infection on infectiousness of SARS-CoV-2 infections

**DOI:** 10.1101/2022.07.05.22277257

**Authors:** Suelen H. Qassim, Mohammad R. Hasan, Patrick Tang, Hiam Chemaitelly, Houssein H. Ayoub, Hadi M. Yassine, Hebah A. Al-Khatib, Maria K. Smatti, Hanan F. Abdul-Rahim, Gheyath K. Nasrallah, Mohamed Ghaith Al-Kuwari, Abdullatif Al-Khal, Peter Coyle, Imtiaz Gillani, Anvar Hassan Kaleeckal, Riyazuddin Mohammad Shaik, Ali Nizar Latif, Einas Al-Kuwari, Andrew Jeremijenko, Adeel A. Butt, Roberto Bertollini, Hamad Eid Al-Romaihi, Mohamed H. Al-Thani, Laith J. Abu-Raddad

## Abstract

In 2021, Qatar experienced considerable incidence of SARS-CoV-2 infection that was dominated sequentially by the Alpha, Beta, and Delta variants. Using the cycle threshold (Ct) value of an RT-qPCR-positive test to proxy the inverse of infectiousness, we investigated infectiousness of SARS-CoV-2 infections by variant, age, sex, vaccination status, prior infection status, and reason for testing in a random sample of 18,355 RT-qPCR-genotyped infections. Regression analyses were conducted to estimate associations with the Ct value of RT-qPCR-positive tests. Compared to Beta infections, Alpha and Delta infections demonstrated 2.56 higher Ct cycles (95% CI: 2.35-2.78), and 4.92 fewer cycles (95% CI: 4.67-5.16), respectively. The Ct value declined gradually with age and was especially high for children <10 years of age, signifying lower infectiousness of small children. Children <10 years of age had 2.18 higher Ct cycles (95% CI: 1.88-2.48) than those 10-19 years of age. Compared to unvaccinated individuals, the Ct value was higher among individuals who had received one or two vaccine doses, but the Ct value decreased gradually with time since the second-dose vaccination. Ct value was 2.07 cycles higher (95% CI: 1.42-2.72) for those with a prior infection than those without prior infection. The Ct value was lowest among individuals tested because of symptoms and was highest among individuals tested as a travel requirement. Delta was substantially more infectious than Beta. Prior immunity, whether due to vaccination or prior infection, is associated with lower infectiousness of breakthrough infections, but infectiousness increases gradually with time since the second-dose vaccination.

## Introduction

The severe acute respiratory syndrome coronavirus 2 (SARS-CoV-2) pandemic continues with progressive viral evolution more than two years after it first emerged.(1) Between January 18, 2021 and May 31, 2021, Qatar experienced a SARS-CoV-2 Alpha(2) (B.1.1.7) variant wave(3) that was immediately followed by a Beta(2) (B.1.351) variant wave.(4) Starting in June 2021, the Delta(2) (B.1.617.2) variant dominated a prolonged low-incidence phase that persisted until November of 2021.(5-7) We investigated the effects of SARS-CoV-2 variant, age, vaccination, and prior infection on infectiousness of SARS-CoV-2 infections.

The real-time (quantitative) reverse transcription-polymerase chain reaction (RT-qPCR) cycle threshold (Ct) value of an RT-qPCR-positive SARS-CoV-2 test is a measure of the inverse of viral load and correlates strongly with culturable virus.(8) Therefore, it can be used to proxy inverse of SARS-CoV-2 infectiousness.(8-13) Higher Ct values signify lower infectiousness.(8-13) We assessed differences in Ct values in a random sample of 18,355 RT-qPCR-genotyped SARS-CoV-2 infections in relation to variant status, age, sex, vaccination status, prior infection status, and reason for testing.

## Methods

### Study population, data sources, and study design

This cross sectional study was conducted in the resident population of Qatar, applying a methodology used recently to investigate effects of the BA.1/BA.2 subvariant, vaccination, and prior infection on infectiousness of SARS-CoV-2 Omicron(2) (B.1.1.529) infections.(14) Similarly, several effects on the infectiousness of SARS-CoV-2 infections were investigated including pre-Omicron variants (Alpha, Beta, and Delta), mRNA COVID-19 vaccination status (BNT162b2(15) and mRNA-1273(16)), prior infection status, reason for RT-qPCR testing, study-period month of the RT-qPCR test (to account for the evolving phase of SARS-CoV-2 incidence), and demographic factors, including sex, age, and nationality.

The present study was conducted on a sample of 18,355 SARS-CoV-2 RT-qPCR-positive swabs that were collected randomly on a weekly basis from among all RT-qPCR-confirmed infections in Qatar between March 23, 2021 and November 6, 2021. These documented infections were RT-qPCR genotyped as part of a national project for surveillance of SARS-CoV-2 variants in Qatar.(7, 17-19) Details of laboratory methods for RT-qPCR testing and variant ascertainment are provided in Section 1 of the Supplementary Material. Coronavirus disease 2019 (COVID-19) laboratory testing, vaccination, clinical infection, and demographic data for this population were extracted from the national, federated SARS-CoV-2 databases, which include all RT-qPCR testing, reason for RT-qPCR testing, COVID-19 vaccinations, and related demographic details since the start of the pandemic. Further description of Qatar’s national COVID-19 databases has been reported previously.(4, 6, 7, 13, 20)

Every SARS-CoV-2 RT-qPCR test conducted in Qatar is classified based on the reason for testing (clinical symptoms, contact tracing, surveys or random testing campaigns, individual requests, routine healthcare testing, pre-travel, at port of entry, or other). RT-qPCR testing is performed at a mass scale and most infections are diagnosed not for appearance of symptoms, but because of routine testing.(6) Qatar has unusually young, diverse demographics, in that only 9% of its residents are ≥50 years of age, and 89% are expatriates from over 150 countries.(20, 21) Nearly all individuals were vaccinated in Qatar; however, vaccinations performed elsewhere were recorded in the health system at the port of entry upon arrival to Qatar, per national requirements.

For standardization of RT-qPCR Ct values, we analyzed only RT-qPCR-confirmed infections diagnosed with the TaqPath COVID-19 Combo Kit (Thermo Fisher Scientific, USA(22)). For each individual, all RT-qPCR-positive swabs during the study period were included, provided that at least 90 days had elapsed between two consecutive positive swabs to avoid inclusion of positive results from the same infectious episode.(23, 24) A summary measure was derived for the primary outcome, the RT-qPCR Ct value,(25) by averaging Ct values of the N, ORF1ab, and S gene targets. This average Ct value was used as the dependent variable in all analyses.

Both vaccination status and prior infection status were ascertained at the time of the RT-qPCR test. Vaccination status was defined by the number of administered vaccine doses and months elapsed since the last vaccine dose, with one month defined as 30 days. Only individuals vaccinated with BNT162b2(15) or mRNA-1273(16) vaccines were included in the analyses, as these have been the vaccines of choice in the COVID-19 immunization program in Qatar.(26-28) Rare occurrences of mixed vaccination regimens were excluded. Nearly all vaccinated persons received their second vaccine dose per protocol(15, 16) within 30 days of first dose. History of prior infection was defined as an RT-qPCR-positive test that occurred ≥90 days before the study RT-qPCR-positive test.(4, 5, 29, 30) An RT-qPCR-positive test that occurred <90 days prior to the study RT-qPCR-positive test was not considered a prior infection, but was considered a category of its own. This is because the prior RT-qPCR-positive test and the study RT-qPCR-positive test may both reflect the same infection.(23, 24) A small number of RT-qPCR tests that had no recorded Ct value were excluded from the analysis, but these constituted only 0. 5% of all RT-qPCR-genotyped infections. Otherwise, data for the remaining study variables were complete.

### Oversight

Hamad Medical Corporation and Weill Cornell Medicine-Qatar Institutional Review Boards approved this retrospective study with a waiver of informed consent. The study was reported following the Strengthening the Reporting of Observational Studies in Epidemiology (STROBE) guidelines. The STROBE checklist is found in Supplementary Table 1.

### Statistical analysis

Frequency distributions and measures of central tendency were used to describe the study population with respect to a priori determined factors. These included SARS-CoV-2 variant, vaccination status (factoring dose number and months since vaccination), prior infection status, reason for RT-qPCR testing, study-period month of the RT-qPCR test, and demographic factors, age, sex, and nationality.

Association of each of these factors with Ct value was assessed using univariable linear regression analyses. Unadjusted β coefficients, 95% confidence intervals (CIs), and the F-test of overall covariate significance were reported. Adjusted β coefficients and associated 95% CIs and p-values were estimated using multivariable linear regression analyses that included all covariates in the model.

Two-sided p-value <0.05 indicated statistical significance. Interactions were not considered. Statistical analyses were conducted in STATA/SE version 16.(31)

## Results

Figure 1 shows the process of selecting the study population and Table 1 describes study population characteristics. This was a national study involving a random sample of 18,355 RT-qPCR-confirmed SARS-CoV-2 infections. Therefore, the study population is broadly representative of the population of Qatar. The sample included 3,347 (18.2%) Alpha infections, 5,576 (30.4%) Beta infections, and 9,432 (51.4%) Delta infections (Table 1).

**Table 1.** Characteristics of the 18,355 RT-qPCR-genotyped SARS-CoV-2 infections between March 23 and November 6, 2021.

| Characteristics | Total sample<br>N (%) | Alpha<br>infections<br>N (%) | Beta<br>infections<br>N (%) | Delta<br>infections<br>N (%) |
| --- | --- | --- | --- | --- |
| **Total N** | 18,355 | 3,347 (18.2) | 5,576 (30.4) | 9,432 (51.4) |
| **Demographic characteristics** |  |  |  |  |
| **Median age (IQR) — years** | 32 (22-41) | 33 (25-41) | 33 (25-42) | 31 (18-40) |
| **Age group in years — no. (%)** |  |  |  |  |
| <10 | 1,956 (10.7) | 286 (8.5) | 424 (7.6) | 1,246 (13.2) |
| 10-19 | 2,042 (11.1) | 277 (8.3) | 499 (8.9) | 1,266 (13.4) |
| 20-29 | 3,670 (20.0) | 749 (22.4) | 1,104 (19.8) | 1,817 (19.3) |
| 30-39 | 5,624 (30.6) | 1,104 (33.0) | 1,866 (33.5) | 2,654 (28.1) |
| 40-49 | 3,263 (17.8) | 627 (18.7) | 1,074 (19.3) | 1,562 (16.6) |
| 50-59 | 1,240 (6.8) | 210 (6.3) | 431 (7.7) | 599 (6.4) |
| 60-69 | 426 (2.3) | 73 (2.2) | 134 (2.4) | 219 (2.3) |
| 70-79 | 95 (0.5) | 14 (0.4) | 31 (0.6) | 50 (0.5) |
| 80+ | 39 (0.2) | 7 (0.2) | 13 (0.2) | 19 (0.2) |
| **Sex** |  |  |  |  |
| Female | 6,805 (37.1) | 1,114 (33.3) | 1,813 (32.5) | 3,878 (41.1) |
| Male | 11,550 (62.9) | 2,233 (66.7) | 3,763 (67.5) | 5,554 (58.9) |
| **Nationality<sup>†</sup>** |  |  |  |  |
| Bangladeshi | 1,077 (5.9) | 240 (7.2) | 470 (8.4) | 367 (3.9) |
| Egyptian | 1,098 (6.0) | 115 (3.4) | 311 (5.6) | 672 (7.1) |
| Filipino | 1,555 (8.5) | 340 (10.2) | 582 (10.4) | 633 (6.7) |
| Indian | 3,873 (21.1) | 855 (25.5) | 1,355 (24.3) | 1,663 (17.6) |
| Nepalese | 1,084 (5.9) | 225 (6.7) | 459 (8.2) | 400 (4.2) |
| Pakistani | 1,017 (5.5) | 223 (6.7) | 279 (5.0) | 515 (5.5) |
| Qatari | 3,903 (21.3) | 555 (16.6) | 850 (15.2) | 2,498 (26.5) |
| Sri Lankan | 488 (2.7) | 106 (3.2) | 216 (3.9) | 166 (1.8) |
| Sudanese | 394 (2.1) | 92 (2.7) | 108 (1.9) | 194 (2.1) |
| Other nationalities<sup>‡</sup> | 3,866 (21.1) | 596 (17.8) | 946 (17.0) | 2,324 (24.6) |
| **RT-qPCR test characteristics** |  |  |  |  |
| **Reason for RT-qPCR testing** |  |  |  |  |
| Clinical suspicion | 6,499 (35.4) | 1,067 (31.9) | 2,477 (44.4) | 2,955 (31.3) |
| Contact tracing | 2,261 (12.3) | 404 (12.1) | 775 (13.9) | 1,082 (11.5) |
| Healthcare routine testing | 532 (2.9) | 115 (3.4) | 218 (3.9) | 199 (2.1) |
| Survey | 2,770 (15.1) | 552 (16.5) | 938 (16.8) | 1,280 (13.6) |
| Port of entry | 3,245 (17.7) | 522 (15.6) | 291 (5.2) | 2,432 (25.8) |
| Pre-travel | 1,243 (6.8) | 201 (6.0) | 225 (4.0) | 817 (8.7) |
| Individual request | 1,620 (8.8) | 435 (13.0) | 603 (10.8) | 582 (6.2) |
| Other | 185 (1.0) | 51 (1.5) | 49 (0.9) | 85 (0.9) |
| **RT-qPCR test study-period month** |  |  |  |  |
| 23 March-21 April, 2021 | 5,185 (28.2) | 1,415 (42.3) | 3,564 (63.9) | 206 (2.2) |
| 22 April-21 May, 2021 | 2,502 (13.6) | 736 (22.0) | 1,262 (22.6) | 504 (5.3) |
| 22 May-20 June, 2021 | 1,287 (7.0) | 597 (17.8) | 226 (4.1) | 464 (4.9) |
| 21 June-20 July, 2021 | 1,263 (6.9) | 258 (7.7) | 111 (2.0) | 894 (9.5) |
| 21 July-19 August, 2021 | 3,647 (19.9) | 253 (7.6) | 326 (5.8) | 3,068 (32.5) |
| 20 August-18 September, 2021 | 2,807 (15.3) | 88 (2.6) | 87 (1.6) | 2,632 (27.9) |
| 19 September-18 October, 2021 | 976 (5.3) | 0 (0.0) | 0 (0.0) | 976 (10.3) |
| 19 October-06 November, 2021 | 688 (3.7) | 0 (0.0) | 0 (0.0) | 688 (7.3) |
| **Vaccine and natural immunity** |  |  |  |  |
| **Vaccination status** |  |  |  |  |
| Unvaccinated | 12,865 (70.1) | 2,760 (82.5) | 4,429 (79.4) | 5,676 (60.2) |
| One dose | 1,053 (5.7) | 239 (7.1) | 582 (10.4) | 232 (2.5) |
| Two doses |  |  |  |  |
| <3 months before the RT-qPCR test | 1,388 (7.6) | 258 (7.7) | 449 (8.1) | 681 (7.2) |
| 3-<6 months before the RT-qPCR test | 2,469 (13.5) | 86 (2.6) | 107 (1.9) | 2,276 (24.1) |
| 6-<9 months before the RT-qPCR test | 567 (3.1) | 3 (0.1) | 7 (0.1) | 557 (5.9) |
| ≥9 months before the RT-qPCR test | 4 (0.0) | 0 (0.0) | 0 (0.0) | 4 (0.0) |
| Three doses |  |  |  |  |
| ≤1 month before the RT-qPCR test | 5 (0.0) | 1 (0.0) | 2 (0.0) | 2 (0.0) |
| >1 month before the RT-qPCR test | 4 (0.0) | 0 (0.0) | 0 (0.0) | 4 (0.0) |
| **Prior SARS-CoV-2 infection** |  |  |  |  |
| Never | 15,785 (86.0) | 2,540 (75.9) | 4,911 (88.1) | 8,334 (88.4) |
| <90 days before the study RT-qPCR test<sup>§</sup> | 2,362 (12.9) | 772 (23.1) | 605 (10.9) | 985 (10.4) |
| Prior infection<sup>¶</sup> | 208 (1.1) | 35 (1.0) | 60 (1.1) | 113 (1.2) |

Abbreviations: IQR, interquartile range; RT-qPCR, real-time reverse-transcription polymerase chain reaction; SARS-CoV-2, severe acute respiratory syndrome coronavirus 2.
<sup>†</sup>Nationalities were chosen to represent the most populous groups in Qatar.
<sup>‡</sup>These comprise 99 other nationalities in Qatar.
<sup>§</sup>An RT-qPCR-positive test that occurred <90 days before the study RT-qPCR-positive test was included as a separate category in the analysis and was not considered a prior infection. This RT-qPCR-positive test and the study RT-qPCR-positive test may both reflect the same prolonged infection.
<sup>¶</sup>Prior infection was defined as an RT-qPCR-positive test that occurred ≥90 days before the RT-qPCR-positive test that is included in the study.

**Figure 1.**
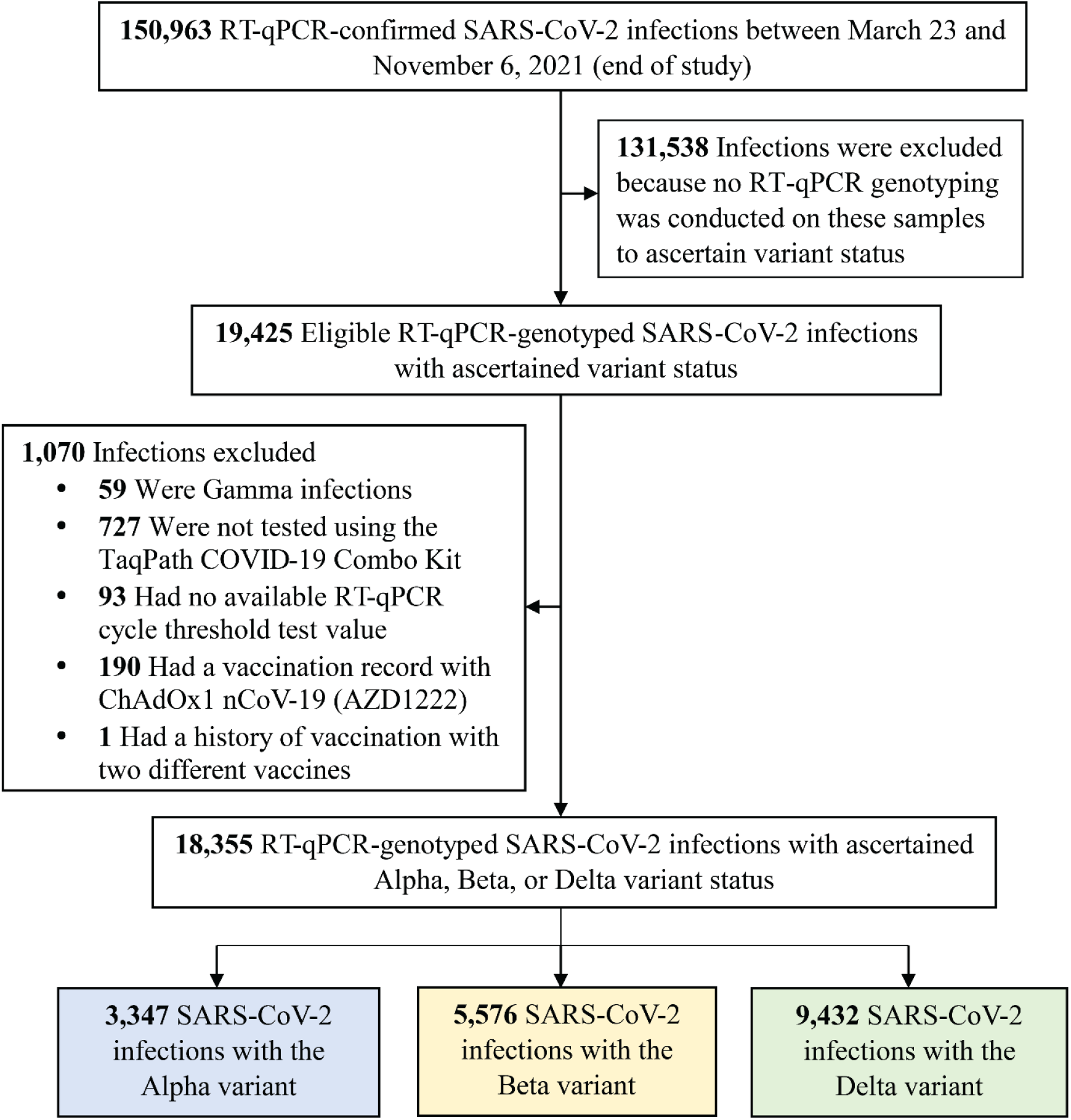
Flowchart describing the population selection process for investigating infectiousness of SARS-CoV-2 Alpha, Beta and Delta infections. Abbreviations: COVID-19, coronavirus disease 2019; RT-qPCR, real-time reverse-transcription polymerase chain reaction; SARS-CoV-2, severe acute respiratory syndrome coronavirus 2.

Compared to Beta infections, Alpha and Delta infections were associated with 2.56 higher Ct cycles (95% CI: 2.35-2.78), and 4.92 fewer cycles (95% CI: 4.67-5.16) (Table 2), respectively, indicating the highest infectiousness for the Delta variant.

**Table 2.**
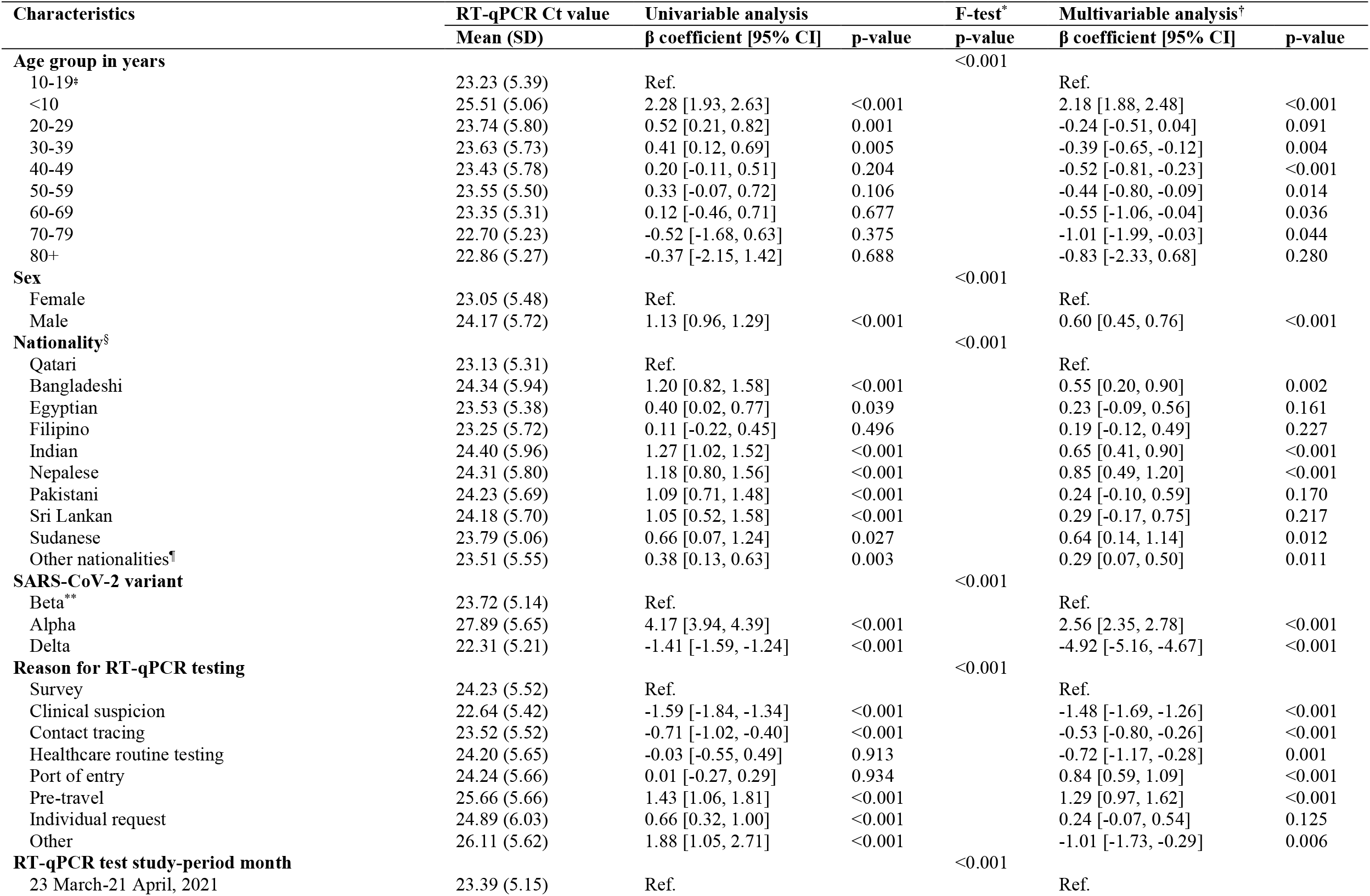
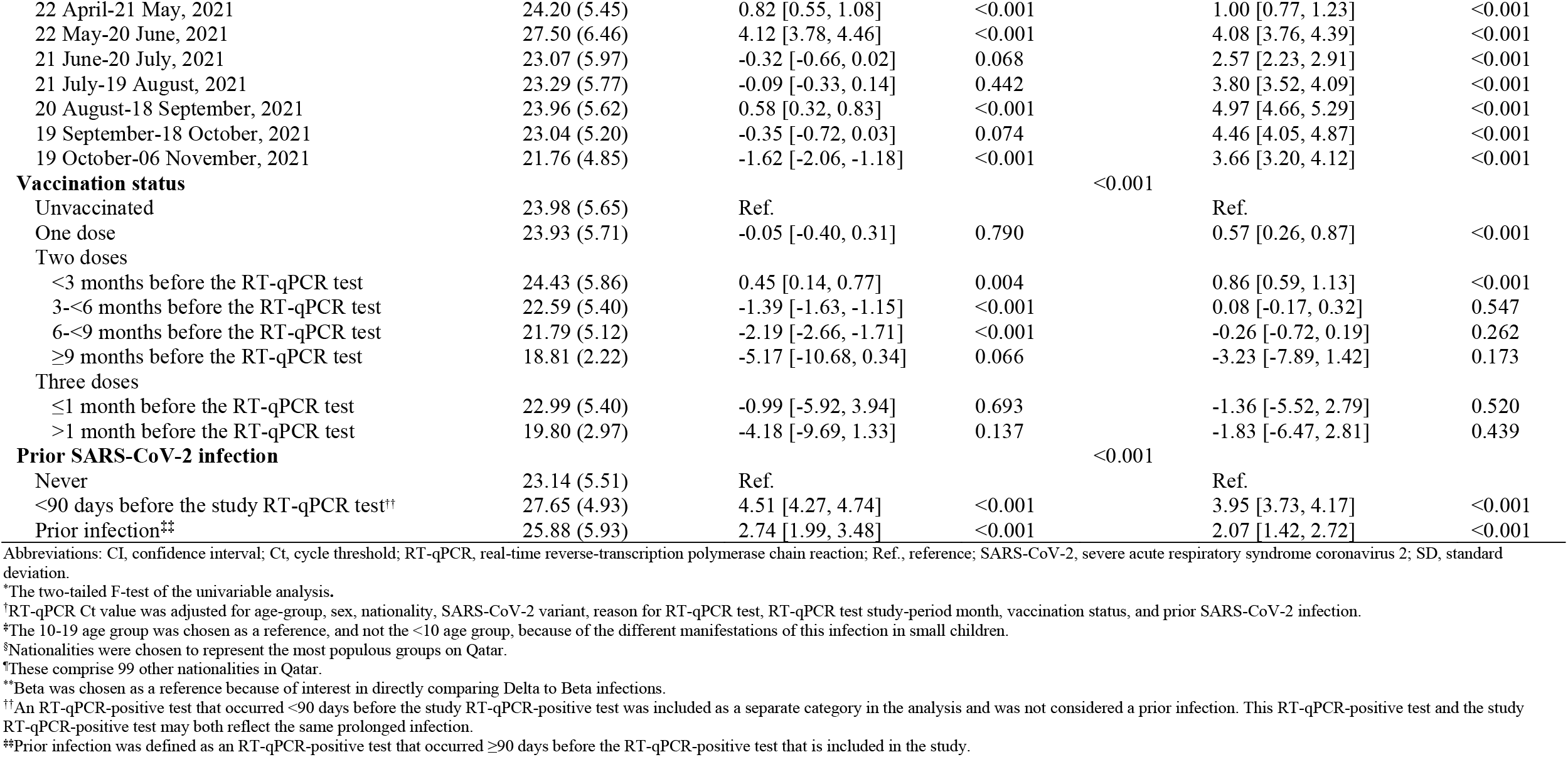
Associations with RT-qPCR Ct value among the 18,355 RT-qPCR-genotyped SARS-CoV-2 infections between March 23 and November 6, 2021.

Ct value declined gradually with age and was especially high for children <10 years of age, signifying lower infectiousness of small children. Children <10 years of age had 2.18 higher Ct cycles (95% CI: 1.88-2.48) than those 10-19 years of age (Table 2). The 10-19 age group was chosen as a reference, and not the <10 age group, because of the different manifestations of this infection in small children. Males had higher Ct values than females and there were some differences in Ct value by nationality.

Compared to unvaccinated individuals, Ct value was higher among individuals who received one or two vaccine doses (Table 2). However, Ct value decreased gradually with time since second-dose vaccination. Very few individuals received a booster dose during the study period (Table 1) to allow for estimation of effect of booster vaccination on Ct value. Ct value was 2.07 cycles higher (95% CI: 1.42-2.72) for those with a prior infection compared to those without prior infection.

The Ct value was lowest when testing was performed due to suspicion of infection exposure (Table 2), such as appearance of symptoms or recent exposure to an infected person (contact tracing). The Ct value was highest for infections diagnosed because of routine testing for reasons unrelated to infection exposure, such as in a random survey or because of travel requirements. Stratified analyses for Alpha (Table 3), Beta (Table 4), and Delta (Table 5) infections suggested similar findings.

**Table 3.**
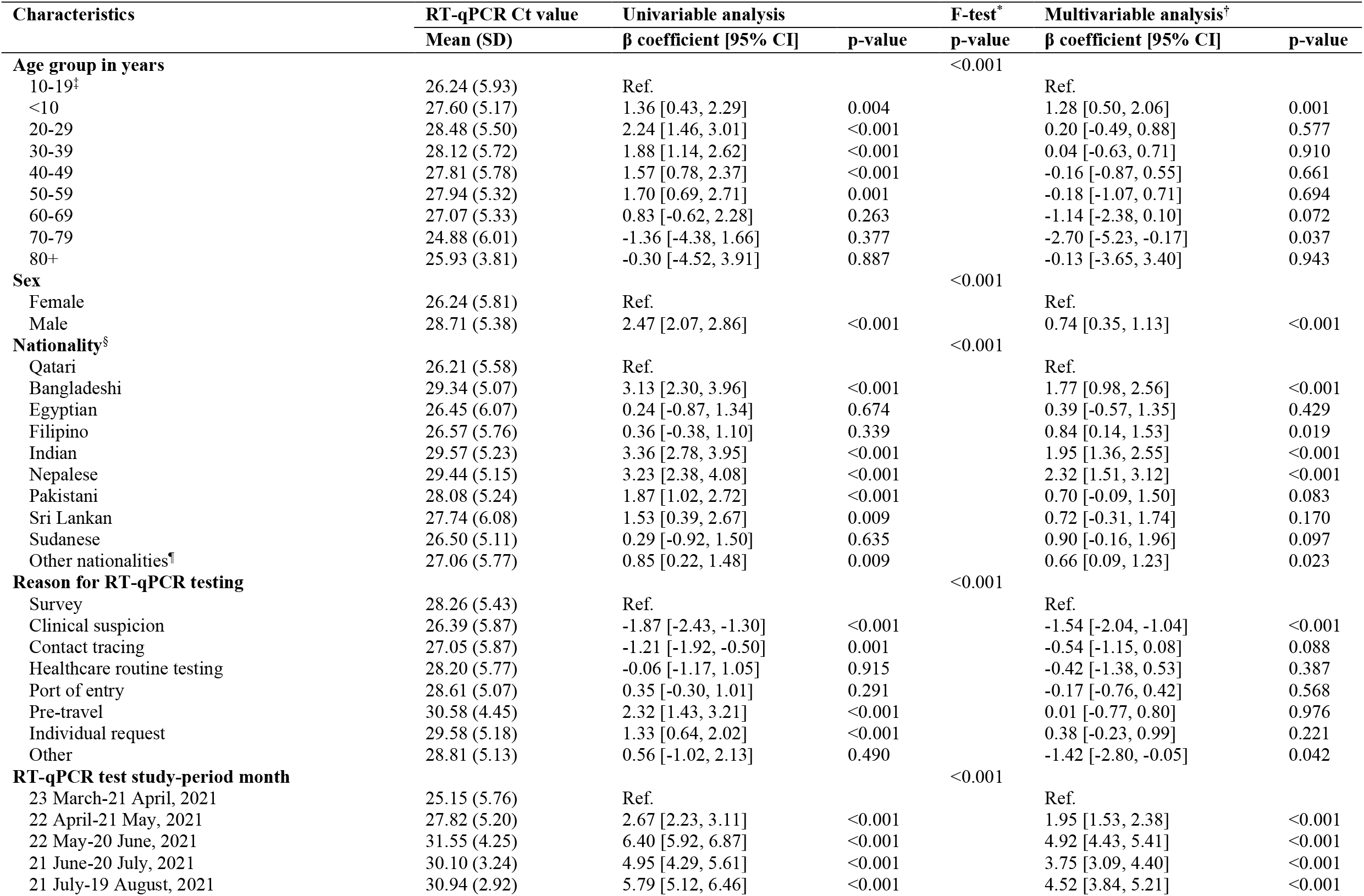
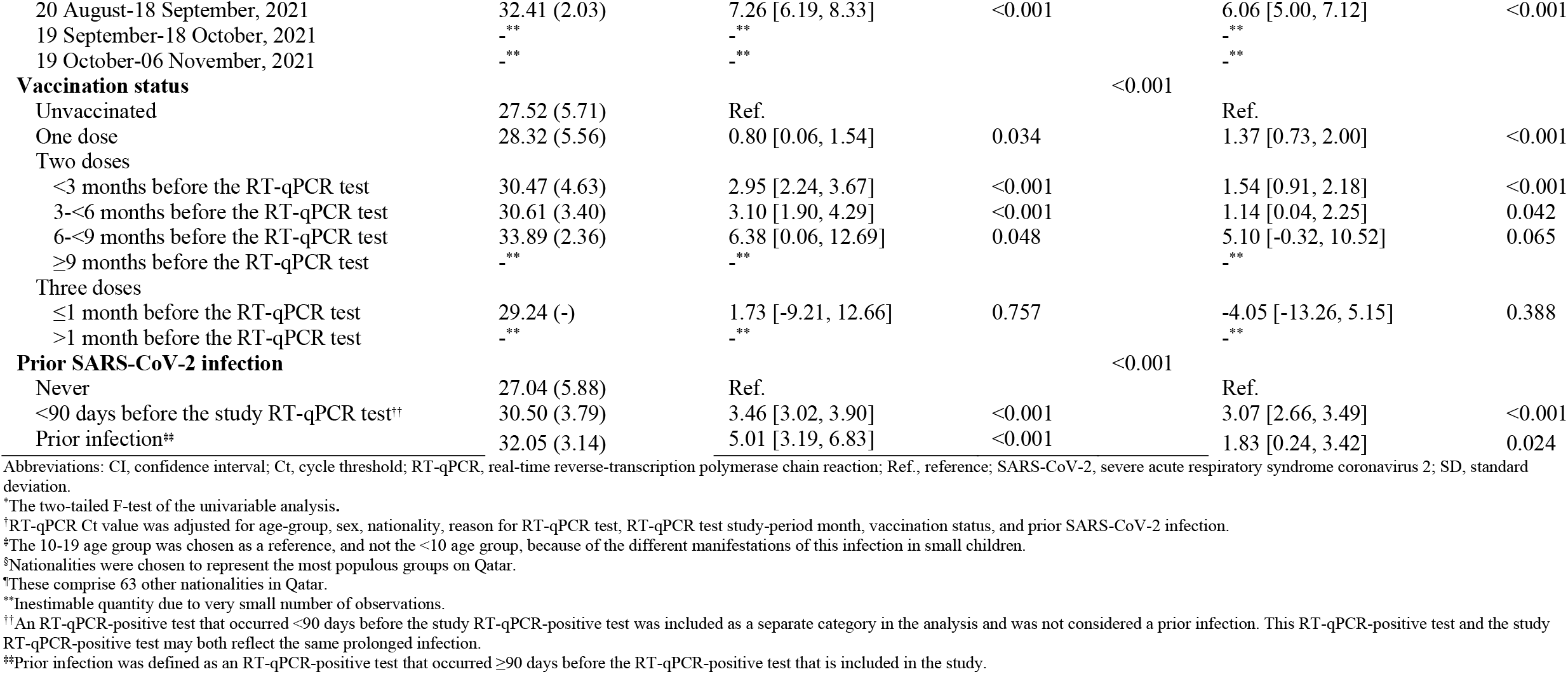
Associations with RT-qPCR Ct value among 3,347 Alpha RT-qPCR-genotyped SARS-CoV-2 infections between March 23 and November 6, 2021.

**Table 4.**
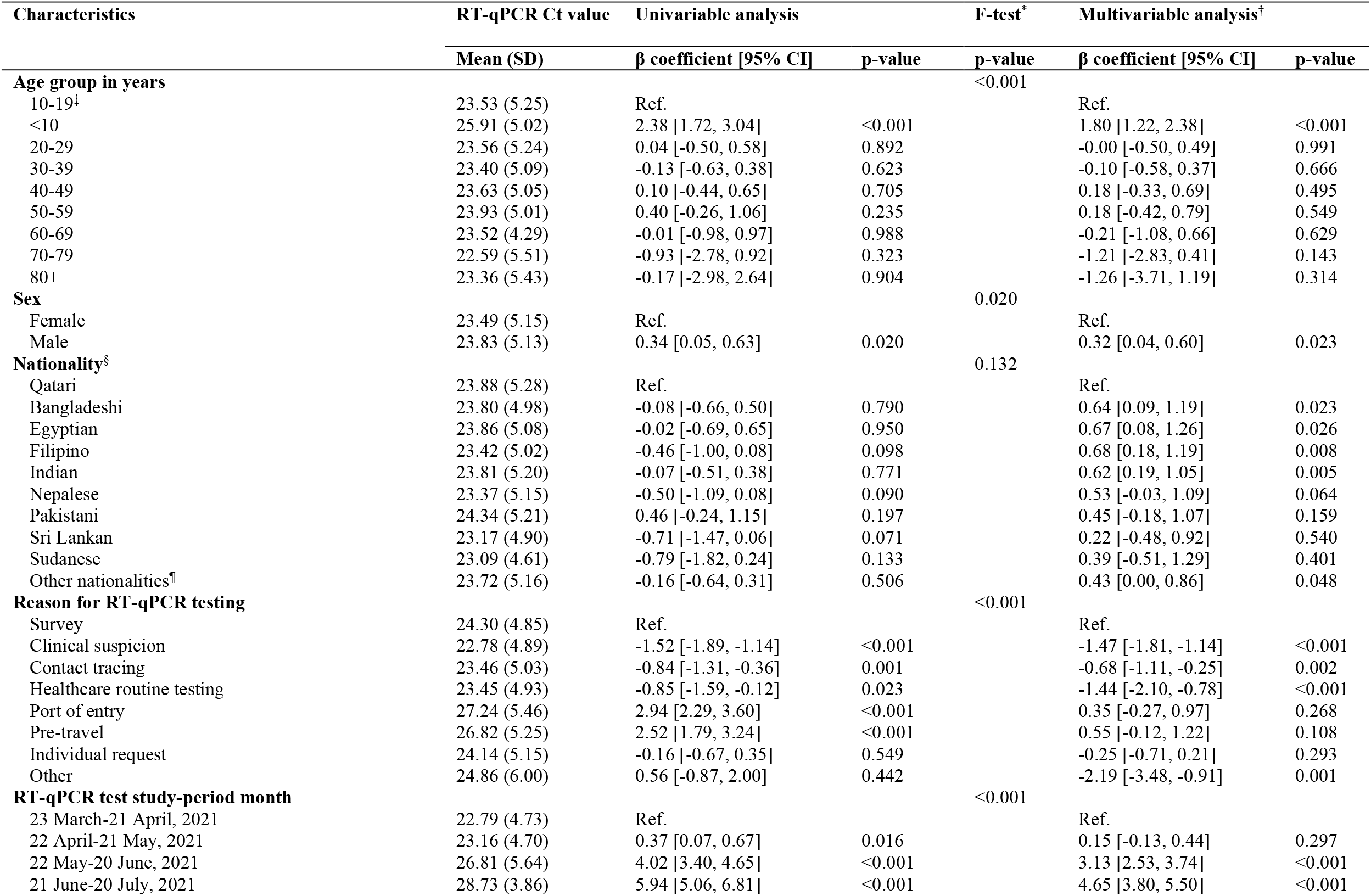
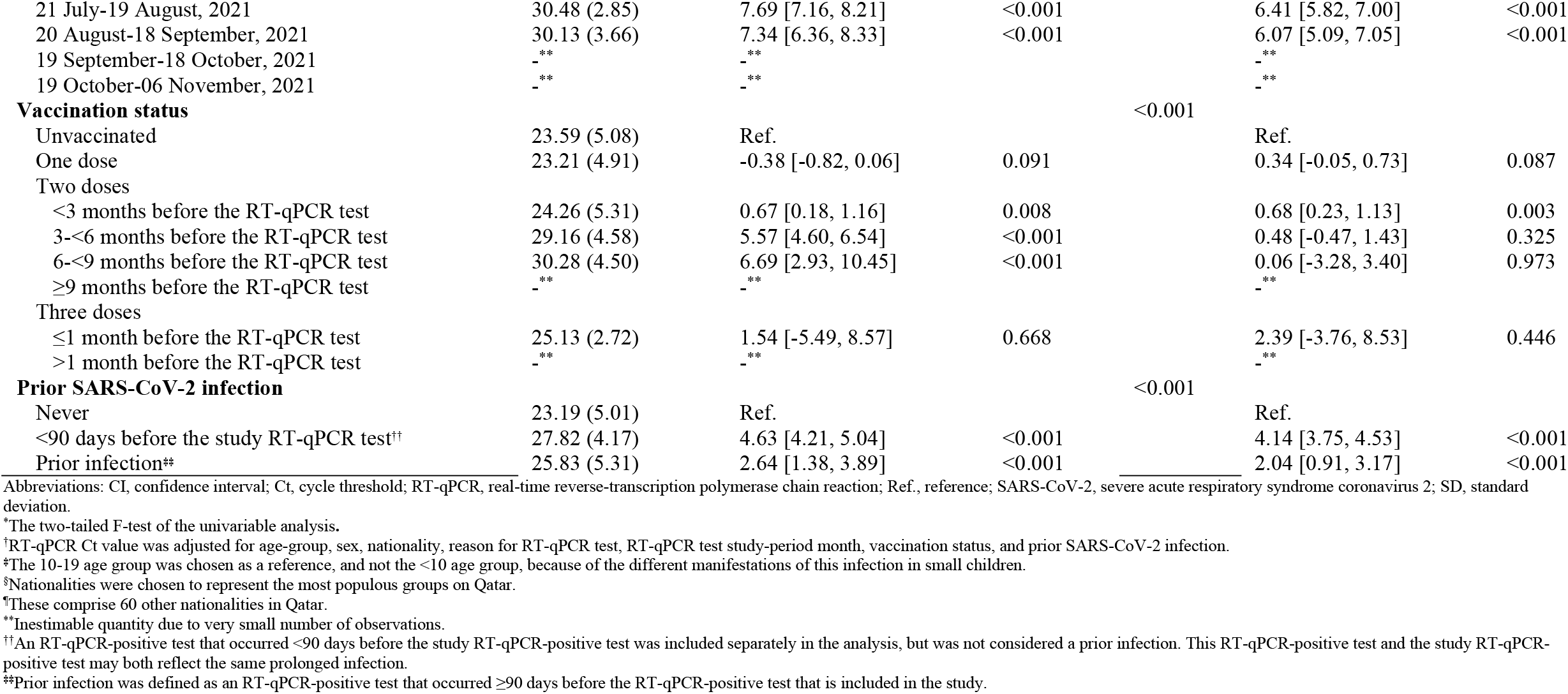
Associations with RT-qPCR Ct value among 5,576 Beta RT-qPCR-genotyped SARS-CoV-2 infections between March 23 and November 6, 2021.

**Table 5.** Associations with RT-qPCR Ct value among 9,432 Delta RT-qPCR-genotyped SARS-CoV-2 infections between March 23 and November 6, 2021.

| Characteristics | RT-qPCR Ct value | Univariable analysis |  | F-test* | Multivariable analysis† |  |
| --- | --- | --- | --- | --- | --- | --- |
| | Mean (SD) | $\beta$ coefficient [95% CI] | p-value | p-value | $\beta$ coefficient [95% CI] | p-value |
| **Age group in years** |  |  |  | <0.001 |  |  |
| 10-19‡ | 22.45 (5.07) | Ref. |  |  | Ref. |  |
| <10 | 24.89 (4.91) | 2.44 [2.04, 2.84] | <0.001 |  | 2.45 [2.07, 2.83] | <0.001 |
| 20-29 | 21.90 (5.12) | -0.55 [-0.91, -0.18] | 0.003 |  | -0.54 [-0.91, -0.16] | 0.005 |
| 30-39 | 21.93 (5.15) | -0.52 [-0.86, -0.18] | 0.003 |  | -0.64 [-1.00, -0.28] | <0.001 |
| 40-49 | 21.53 (5.24) | -0.92 [-1.30, -0.54] | <0.001 |  | -1.00 [-1.39, -0.61] | <0.001 |
| 50-59 | 21.75 (4.95) | -0.70 [-1.20, -0.20] | 0.006 |  | -0.84 [-1.34, -0.34] | 0.001 |
| 60-69 | 22.01 (5.28) | -0.44 [-1.17, 0.29] | 0.239 |  | -0.61 [-1.33, 0.10] | 0.093 |
| 70-79 | 22.16 (4.76) | -0.29 [-1.73, 1.16] | 0.696 |  | -0.04 [-1.42, 1.33] | 0.952 |
| 80+ | 21.39 (5.29) | -1.06 [-3.37, 1.26] | 0.371 |  | -0.76 [-2.94, 1.42] | 0.493 |
| **Sex** |  |  |  | <0.001 |  |  |
| Female | 21.92 (5.12) | Ref. |  |  | Ref. |  |
| Male | 22.58 (5.26) | 0.66 [0.44, 0.87] | <0.001 |  | 0.48 [0.27, 0.70] | <0.001 |
| **Nationality§** |  |  |  | <0.001 |  |  |
| Qatari | 22.19 (4.95) | Ref. |  |  | Ref. |  |
| Bangladeshi | 21.75 (5.63) | -0.44 [-1.01, 0.13] | 0.129 |  | -0.14 [-0.71, 0.42] | 0.617 |
| Egyptian | 22.88 (5.22) | 0.68 [0.24, 1.13] | 0.002 |  | -0.08 [-0.50, 0.35] | 0.717 |
| Filipino | 21.30 (5.46) | -0.89 [-1.35, -0.44] | <0.001 |  | -0.37 [-0.83, 0.08] | 0.106 |
| Indian | 22.21 (5.27) | 0.02 [-0.30, 0.34] | 0.900 |  | -0.29 [-0.64, 0.06] | 0.110 |
| Nepalese | 22.50 (5.17) | 0.31 [-0.24, 0.86] | 0.265 |  | -0.12 [-0.68, 0.44] | 0.677 |
| Pakistani | 22.50 (5.29) | 0.30 [-0.19, 0.80] | 0.227 |  | 0.07 [-0.41, 0.56] | 0.766 |
| Sri Lankan | 23.21 (5.54) | 1.02 [0.20, 1.83] | 0.015 |  | 0.33 [-0.44, 1.10] | 0.399 |
| Sudanese | 22.89 (4.85) | 0.70 [-0.06, 1.46] | 0.073 |  | 0.87 [0.16, 1.58] | 0.017 |
| Other nationalities¶ | 22.51 (5.25) | 0.32 [0.02, 0.61] | 0.034 |  | 0.13 [-0.15, 0.42] | 0.357 |
| **Reason for RT-qPCR testing** |  |  |  | <0.001 |  |  |
| Survey | 22.44 (5.08) | Ref. |  |  | Ref. |  |
| Clinical suspicion | 21.16 (4.99) | -1.28 [-1.61, -0.94] | <0.001 |  | -1.36 [-1.69, -1.04] | <0.001 |
| Contact tracing | 22.25 (5.15) | -0.19 [-0.60, 0.23] | 0.371 |  | -0.37 [-0.76, 0.03] | 0.073 |
| Healthcare routine testing | 22.71 (5.26) | 0.27 [-0.49, 1.04] | 0.483 |  | -0.17 [-0.90, 0.56] | 0.650 |
| Port of entry | 22.94 (5.18) | 0.50 [0.16, 0.85] | 0.004 |  | 0.93 [0.59, 1.27] | <0.001 |
| Pre-travel | 24.14 (5.27) | 1.70 [1.25, 2.15] | <0.001 |  | 1.80 [1.37, 2.22] | <0.001 |
| Individual request | 22.15 (5.41) | -0.29 [-0.79, 0.21] | 0.262 |  | 0.36 [-0.14, 0.86] | 0.158 |
| Other | 25.20 (5.18) | 2.76 [1.63, 3.89] | <0.001 |  | -0.01 [-1.11, 1.09] | 0.989 |
| **RT-qPCR test study-period month** |  |  |  | <0.001 |  |  |
| 23 March-21 April, 2021 | 21.55 (4.55) | Ref. |  |  | Ref. |  |
| 22 April-21 May, 2021 | 21.54 (4.89) | -0.02 [-0.85, 0.81] | 0.968 |  | -0.17 [-0.96, 0.61] | 0.666 |
| 22 May-20 June, 2021 | 22.64 (5.69) | 1.09 [0.25, 1.93] | 0.011 |  | 1.05 [0.24, 1.86] | 0.011 |
| 21 June-20 July, 2021 | 20.34 (4.44) | -1.22 [-1.99, -0.44] | 0.002 |  | -1.14 [-1.90, -0.38] | 0.003 |
| 21 July-19 August, 2021 | 21.90 (5.07) | 0.35 [-0.38, 1.07] | 0.348 |  | 0.13 [-0.58, 0.84] | 0.722 |
| 20 August-18 September, 2021 | 23.47 (5.41) | 1.92 [1.19, 2.65] | <0.001 |  | 1.55 [0.84, 2.27] | <0.001 |
| 19 September-18 October, 2021 | 23.04 (5.20) | 1.49 [0.72, 2.26] | <0.001 |  | 1.23 [0.47, 1.98] | 0.001 |
| 19 October-06 November, 2021 | 21.76 (4.85) | 0.21 [-0.59, 1.01] | 0.606 |  | 0.26 [-0.53, 1.05] | 0.524 |
| **Vaccination status** |  |  |  | <0.001 |  |  |
| Unvaccinated | 22.56 (5.31) | Ref. |  |  | Ref. |  |
| One dose | 21.22 (5.24) | -1.34 [-2.02, -0.66] | <0.001 |  | 0.35 [-0.29, 0.99] | 0.281 |
| Two doses |  |  |  |  |  |  |
| <3 months before the RT-qPCR test | 22.26 (4.98) | -0.30 [-0.72, 0.11] | 0.151 |  | 0.87 [0.47, 1.27] | <0.001 |
| 3-<6 months before the RT-qPCR test | 21.98 (5.04) | -0.59 [-0.84, -0.33] | <0.001 |  | 0.22 [-0.06, 0.50] | 0.120 |
| 6-<9 months before the RT-qPCR test | 21.62 (4.97) | -0.94 [-1.40, -0.49] | <0.001 |  | -0.23 [-0.72, 0.25] | 0.347 |
| ≥9 months before the RT-qPCR test | 18.81 (2.22) | -3.75 [-8.85, 1.35] | 0.149 |  | -3.15 [-7.87, 1.57] | 0.191 |
| Three doses |  |  |  |  |  |  |
| ≤1 month before the RT-qPCR test | 17.72 (2.35) | -4.84 [-12.05, 2.36] | 0.188 |  | -3.46 [-10.16, 3.23] | 0.311 |
| >1 month before the RT-qPCR test | 19.80 (2.97) | -2.77 [-7.87, 2.33] | 0.287 |  | -1.18 [-5.89, 3.53] | 0.623 |
| **Prior SARS-CoV-2 infection** |  |  |  | <0.001 |  |  |
| Never | 21.93 (5.11) | Ref. |  |  | Ref. |  |
| <90 days before the study RT-qPCR test<sup>††</sup> | 25.32 (4.95) | 3.39 [3.05, 3.72] | <0.001 |  | 4.03 [3.69, 4.38] | <0.001 |
| Prior infection<sup>‡‡</sup> | 24.00 (5.64) | 2.07 [1.13, 3.02] | <0.001 |  | 2.15 [1.26, 3.04] | <0.001 |

Abbreviations: CI, confidence interval; Ct, cycle threshold; RT-qPCR, real-time reverse-transcription polymerase chain reaction; Ref., reference; SARS-CoV-2, severe acute respiratory syndrome coronavirus 2; SD, standard deviation.
<sup>†</sup>The two-tailed F-test of the univariable analysis.
<sup>††</sup>RT-qPCR Ct value was adjusted for age-group, sex, nationality, reason for RT-qPCR test, RT-qPCR test study-period month, vaccination status, and prior SARS-CoV-2 infection.
<sup>‡</sup>The 10-19 age group was chosen as a reference, and not the <10 age group, because of the different manifestations of this infection in small children.
<sup>‡‡</sup>Nationalities were chosen to represent the most populous groups on Qatar.
<sup>†††</sup>These comprise 87 other nationalities in Qatar.
<sup>‡‡‡</sup>Inestimable quantity due to very small number of observations.
<sup>††††</sup>An RT-qPCR-positive test that occurred <90 days before the study RT-qPCR-positive test was included as a separate category in the analysis and was not considered a prior infection. This RT-qPCR-positive test and the study RT-qPCR-positive test may both reflect the same prolonged infection.
<sup>‡‡‡‡</sup>Prior infection was defined as an RT-qPCR-positive test that occurred ≥90 days before the RT-qPCR-positive test that is included in the study.

## Discussion

Delta infections were associated with considerably lower Ct values than Beta infections, indicating higher infectiousness of this variant, perhaps because of higher viral load and/or longer duration of infection. This appears to be the first direct comparison of the infectiousness of Delta versus Beta infections, and supports the high infectiousness of the Delta variant compared to pre-Omicron variants such as Alpha, as reported previously.(32, 33) This difference in viral load between Delta and Beta appears also to extend to severity of infection, as Delta infections were found associated with higher severity than Beta infections.(34) Of note that Beta infections were also found earlier to be associated with higher severity than Alpha infections.(35)

Prior immunity against SARS-CoV-2 infection, whether due to vaccination or prior infection, was associated with higher Ct value at infection, and thus lower infectiousness of breakthrough infections. This confirms earlier findings,(13, 14) and suggests that strength of immunity is manifest not only in protection against infection, but also against the infectiousness, if a breakthrough infection occurs.(13) However, this effect appeared to depend on the time since the prior immunological event (Table 2). Ct values decreased gradually with time since second-dose vaccination, paralleling the established pattern of waning of vaccine effectiveness after the second dose.(6, 36, 37)

Ct values decreased with age, perhaps reflecting slower virus clearance with aging (38) and confirming our earlier findings.(14) Ct values were particularly high for infections among small children <10 years of age. This finding supports a lesser role for small children than adults in the transmission of infection, as suggested in studies of secondary transmission within households.(39-41) There were differences in Ct value by sex and nationality, but these may be a consequence of different test-seeking behaviors for different socio-economic groups in Qatar’s diverse population. Ct values also varied by reason for testing, with lower Ct values of infections diagnosed because of suspicion of infection, and higher Ct values of infections diagnosed through routine testing unrelated to infection exposure. This finding also confirms our earlier finding for Omicron infections.(14)

The study has limitations. While both Delta and Beta infections had lower Ct values than Alpha infections, this finding should be interpreted with caution. RT-qPCR genotyping in Qatar started only after the Alpha wave peaked in the first week of March 2021. Many Alpha infections may have been older or prolonged infections, rather than recent infections, explaining the higher Ct values of these infections.

Eligible individuals were selected from the pool of those who had documented RT-qPCR-confirmed infections, but other infections may have occurred that were never documented. It is possible that infections in those with prior infections or those vaccinated are less likely to be diagnosed, perhaps because of minimal or no symptoms.(13) Nevertheless, RT-qPCR testing in Qatar is done at a mass scale, where a significant proportion of the population is being tested every week.(6) The majority of infections are identified not because of symptoms, but because of routine testing for other reasons (Table 1).(6) The date of symptom onset was not available for symptomatic cases. Therefore, an analysis factoring the duration between symptom onset and RT-qPCR test was not possible.

A small number of RT-qPCR tests had missing Ct values and were excluded from the analysis, but these constituted only 0.5% of all RT-qPCR-genotyped infections. The study population consisted mostly of working-age adults; thus, the results may not be generalizable to the elderly. Too few individuals received the booster dose during the study period to allow for estimation of effect of booster vaccination on Ct values.

In conclusion, the Delta variant appears substantially more infectious than the Beta variant, explaining its global reach in the pre-Omicron era. Infectiousness of SARS-CoV-2 infections increases with age, apparently reflecting slower virus clearance with aging. Prior immunity against SARS-CoV-2 infection, whether due to vaccination or prior infection, is associated with lower infectiousness of breakthrough infections. However, infectiousness of breakthrough infections increases gradually with time since second-dose vaccination, paralleling the waning of vaccine effectiveness after the second dose.

## Data Availability

The dataset of this study is a property of the Qatar Ministry of Public Health that was provided to the researchers through a restricted-access agreement that prevents sharing the dataset with a third party or publicly. Future access to this dataset can be considered through a direct application for data access to Her Excellency the Minister of Public Health (https://www.moph.gov.qa/english/Pages/default.aspx). Aggregate data are available within the manuscript and its Supplementary information.

## Acknowledgements

We acknowledge the many dedicated individuals at Hamad Medical Corporation, the Ministry of Public Health, the Primary Health Care Corporation, Qatar Biobank, Sidra Medicine, and Weill Cornell Medicine-Qatar for their diligent efforts and contributions to make this study possible. The authors are grateful for institutional salary support from the Biomedical Research Program and the Biostatistics, Epidemiology, and Biomathematics Research Core, both at Weill Cornell Medicine-Qatar, as well as for institutional salary support provided by the Ministry of Public Health, Hamad Medical Corporation, and Sidra Medicine. The authors are also grateful for the Qatar Genome Programme and Qatar University Biomedical Research Center for institutional support for the reagents needed for the viral genome sequencing. The funders of the study had no role in study design, data collection, data analysis, data interpretation, or writing of the article. Statements made herein are solely the responsibility of the authors.

## Author contributions

SHQ co-designed the study, performed the statistical analyses, and co-wrote the first draft of the article. PT and MRH conducted the multiplex, RT-qPCR variant screening and viral genome sequencing. HC co-designed the study and supported the statistical analyses. LJA conceived and co-designed the study, led the statistical analyses, and co-wrote the first draft of the article. All authors contributed to data collection and acquisition, database development, discussion and interpretation of the results, and to the writing of the manuscript. All authors have read and approved the final manuscript.

## Supplementary Material

### 1. Laboratory methods and variant ascertainment

#### 1.1 Real-time reverse-transcription polymerase chain reaction testing

Nasopharyngeal and/or oropharyngeal swabs were collected for polymerase chain reaction (PCR) testing and placed in Universal Transport Medium (UTM). Aliquots of UTM were: 1) extracted on KingFisher Flex (Thermo Fisher Scientific, USA), MGISP-960 (MGI, China), or ExiPrep 96 Lite (Bioneer, South Korea) followed by testing with real-time reverse-transcription PCR (RT-qPCR) using TaqPath COVID-19 Combo Kits (Thermo Fisher Scientific, USA) on an ABI 7500 FAST (Thermo Fisher Scientific, USA); 2) tested directly on the Cepheid GeneXpert system using the Xpert Xpress SARS-CoV-2 (Cepheid, USA); or 3) loaded directly into a Roche cobas 6800 system and assayed with the cobas SARS-CoV-2 Test (Roche, Switzerland). The first assay targets the viral S, N, and ORF1ab gene regions. The second targets the viral N and E-gene regions, and the third targets the ORF1ab and E-gene regions.

All PCR testing was conducted at the Hamad Medical Corporation Central Laboratory or Sidra Medicine Laboratory, following standardized protocols. All PCR testing was performed with extensively used, investigated, and validated commercial platforms having essentially 100% sensitivity and specificity.

#### 1.2 Classification of infections by variant type

Surveillance for SARS-CoV-2 variants in Qatar is mainly based on viral genome sequencing and multiplex RT-qPCR variant screening(1) of random positive clinical samples,(2-7) complemented by deep sequencing of wastewater samples.(4, 8)

The accuracy of the RT-qPCR genotyping was verified against either Sanger sequencing of the receptor-binding domain (RBD) of SARS-CoV-2 surface glycoprotein (S) gene, or by viral whole-genome sequencing on a Nanopore GridION sequencing device. From 236 random samples (27 Alpha-like, 186 Beta-like, and 23 “other” variants), PCR genotyping results for Alpha-like, Beta-like, and ‘other’ variants were in 88.8% (23 out of 27), 99.5% (185 out of 186), and 100% (23 out of 23) agreement with the SARS-CoV-2 lineages assigned by sequencing.

Within the “other” variant category, Sanger sequencing and/or Illumina sequencing of the RBD of SARS-CoV-2 spike gene on 728 random samples confirmed that 701 (96.3%) were Delta cases and 17 (2.3%) were other variant cases, with 10 (1.4%) samples failing lineage assignment.^6,8^ Accordingly, a Delta case was proxied as any “other” case identified through the RT-qPCR based variant screening.

All the variant RT-qPCR screening was conducted at the Sidra Medicine Laboratory following standardized protocols.

## 2. Supplementary tables

**Supplementary table 1.** STROBE checklist for cross-sectional studies.

|  | Item No | Recommendations | Main text page No |
| --- | --- | --- | --- |
| Title and abstract | 1 | (a) Indicate the study’s design with a commonly used term in the title or the abstract | Methods (‘Study population, data sources, and study design’, paragraph 1) |
|  |  | (b) Provide in the abstract an informative and balanced summary of what was done and what was found | Abstract |
| Introduction |  |  |  |
| Background/rationale | 2 | Explain the scientific background and rationale for the investigation being reported | Introduction |
| Objectives | 3 | State specific objectives, including any prespecified hypotheses | Methods (‘Study population, data sources, and study design’) |
| Methods |  |  |  |
| Study design | 4 | Present key elements of study design early in the paper | Methods (‘Study population, data sources, and study design’) |
| Setting | 5 | Describe the setting, locations, and relevant dates, including periods of recruitment, exposure, follow-up, and data collection | Methods (‘Study population, data sources, and study design’) and Section 1 (‘Classification of infections by variant type’) of Supplementary Material |
| Participants | 6 | (a) Give the eligibility criteria, and the sources and methods of selection of participants | Methods (‘Study population, data sources, and study design’) & Figure 1 |
| Variables | 7 | Clearly define all outcomes, exposures, predictors, potential confounders, and effect modifiers. Give diagnostic criteria, if applicable | Methods (‘Study population, data sources, and study design’ & ‘Statistical analysis’) & Sections 1 (‘Classification of infections by variant type’) of Supplementary Material |
| Data sources/measurement | 8 | For each variable of interest, give sources of data and details of methods of assessment (measurement). Describe comparability of assessment methods if there is more than one group | Methods (‘Study population, data sources, and study design’), Table 1, & Section 1 of Supplementary Material |
| Bias | 9 | Describe any efforts to address potential sources of bias | Methods (‘Study population, data sources, and study design’ & ‘Statistical analysis’) |
| Study size | 10 | Explain how the study size was arrived at | Methods (‘Study population, data sources, and study design’) & Figure 1 |
| Quantitative variables | 11 | Explain how quantitative variables were handled in the analyses. If applicable, describe which groupings were chosen and why | Methods (‘Study population, data sources, and study design’ & ‘Statistical analysis’), & Tables 1-5 |
| Statistical methods | 12 | (a) Describe all statistical methods, including those used to control for confounding | Methods (‘Statistical analysis’) |
|  |  | (b) Describe any methods used to examine subgroups and interactions | Methods (‘Study population, data sources, and study design’ & ‘Statistical analysis’), & Tables 3-5 |
|  |  | (c) Explain how missing data were addressed | Methods (‘Study population, data sources, and study design’) & Figure 1 |
|  |  | (d) If applicable, describe analytical methods taking account of sampling strategy | NA |
|  |  | (e) Describe any sensitivity analyses | NA |
| Results |  |  |  |
| Participants | 13 | (a) Report numbers of individuals at each stage of study—eg numbers potentially eligible, examined for eligibility, confirmed eligible, included in the study, completing follow-up, and analysed<br>(b) Give reasons for non-participation at each stage<br>(c) Consider use of a flow diagram | Figure 1 |
| Descriptive data | 14 | (a) Give characteristics of study participants (eg demographic, clinical, social) and information on exposures and potential confounders | Table 1 |
|  |  | (b) Indicate number of participants with missing data for each variable of interest | Methods ('Study population, data sources, and study design') & Figure 1 |
| Outcome data | 15 | Report numbers of outcome events or summary measures | Results & Table 2 |
| Main results | 16 | (a) Give unadjusted estimates and, if applicable, confounder-adjusted estimates and their precision (eg, 95% confidence interval). Make clear which confounders were adjusted for and why they were included | Results & Table 2 |
|  |  | (b) Report category boundaries when continuous variables were categorized | Tables 1-2 |
|  |  | (c) If relevant, consider translating estimates of relative risk into absolute risk for a meaningful time period | NA |
| Other analyses | 17 | Report other analyses done—eg analyses of subgroups and interactions, and sensitivity analyses | Results & Tables 3-5 |
| **Discussion** |  |  |  |
| Key results | 18 | Summarise key results with reference to study objectives | Discussion, paragraph 1 |
| Limitations | 19 | Discuss limitations of the study, taking into account sources of potential bias or imprecision. Discuss both direction and magnitude of any potential bias | Discussion, paragraph 5-6 |
| Interpretation | 20 | Give a cautious overall interpretation of results considering objectives, limitations, multiplicity of analyses, results from similar studies, and other relevant evidence | Discussion, paragraph 1 |
| Generalisability | 21 | Discuss the generalisability (external validity) of the study results | Discussion, paragraph 6 |
| **Other information** |  |  |  |
| Funding | 22 | Give the source of funding and the role of the funders for the present study and, if applicable, for the original study on which the present article is based | Acknowledgements |

Abbreviations: NA: not applicable.

